# Lipid-Modulating Agents for Prevention or Treatment of COVID-19 in Randomized Trials

**DOI:** 10.1101/2021.05.03.21256468

**Authors:** Azita H. Talasaz, Parham Sadeghipour, Maryam Aghakouchakzadeh, Isaac Dreyfus, Hessam Kakavand, Hamid Ariannejad, Aakriti Gupta, Mahesh V. Madhavan, Benjamin W. Van Tassell, David Jimenez, Manuel Monreal, Muthiah Vaduganathan, John Fanikos, Dave L Dixon, Gregory Piazza, Sahil A. Parikh, Deepak L. Bhatt, Gregory YH Lip, Gregg W. Stone, Harlan M. Krumholz, Peter Libby, Samuel Z. Goldhaber, Behnood Bikdeli

**Affiliations:** Department of Clinical Pharmacy, Faculty of Pharmacy, Tehran University of Medical Sciences, Tehran, Iran; Tehran Heart Center, Tehran University of Medical Sciences, Tehran, Iran; Department of Pharmacotherapy and Outcome Science, School of Pharmacy, Virginia Commonwealth University, Richmond, Virginia, USA; Cardiovascular Intervention Research Center, Rajaie Cardiovascular Medical and Research Center, Iran University of Medical Sciences, Tehran, Iran; NewYork-Presbyterian Hospital/Columbia University Irving Medical Center, New York, New York; Clinical Trials Center, Cardiovascular Research Foundation, New York, NY, USA; Center for Outcomes Research and Evaluation (CORE), Yale School of Medicine, New Haven, CT, USA; Pauley Heart Center, Division of Cardiology, Department of Internal Medicine, School of Pharmacy, Virginia Commonwealth University, Richmond, Virginia, USA; Respiratory Department, Hospital Ramón y Cajal and Medicine Department, Universidad de Alcalá (Instituto de Ramón y Cajal de Investigación Sanitaria), Centro de Investigación Biomédica en Red de Enfermedades Respiratorias, Madrid, Spain; Department of Internal Medicine, Hospital Universitari Germans Trials i Pujol, Universidad Católica San Antonio de Murcia, Barcelona, Spain; Division of cardiovascular Medicine Division, Brigham and Women’s Hospital, Harvard Medical School, Boston, MA, USA; Department of Pharmacy, Brigham and Women’s Hospital, Harvard Medical School, Boston, MA, USA; Liverpool Centre for Cardiovascular Science, Liverpool Heart and Chest Hospital, University of Liverpool, Liverpool, United Kingdom; Department of Clinical Medicine, Aalborg University, Aalborg, Denmark; Zena and Michael A. Wiener Cardiovascular Institute, Icahn School of Medicine at Mount Sinai, New York, NY, USA; Department of Health Policy and Administration, Yale School of Public Health, New Haven, CT, USA; Section of Cardiovascular Medicine, Department of Internal Medicine, Yale School of Medicine, New Haven, CT, USA

**Keywords:** COVID-19, lipid modulating agent, statin, fibrate, omega3, niacin

## Abstract

Coronavirus disease 2019 (COVID-19) is associated with systemic inflammation, endothelial activation, and multi-organ manifestations. Lipid modulating agents may be useful in treating patients with COVID-19. They may inhibit viral entry by lipid raft disruption or ameliorate the inflammatory response and endothelial activation. In addition, dyslipidemia with lower high-density lipoprotein cholesterol and higher triglycerides portends worse outcome in patients with COVID-19. Upon a systematic search, 40 RCTs with lipid modulating agents were identified, including 17 statin trials, 14 omega-3 fatty acids RCTs, 3 fibrates RCTs, 5 niacin RCTs, and 1 dalcetrapib RCT for management or prevention of COVID-19. This manuscript summarizes the ongoing or completed randomized controlled trials (RCTs) of lipid modulating agents in COVID-19 and the implications of these trials for patient management.

## INTRODUCTION

Severe acute respiratory syndrome coronavirus 2 (SARS-CoV-2) cellular entry is mediated by attachment to angiotensin converting enzyme 2 (ACE2). Lipid rafts – plasma membrane microdomains mainly composed of cholesterol, glycosphingolipids, and phospholipids, capable of changing their composition in response to stimuli – may play a critical role in this process (1). SARS-CoV-2 can trigger an uncontrolled innate inflammatory response (cytokine storm) leading to local and systemic tissue damage commonly seen in advanced coronavirus disease 2019 (COVID-19) (2). Inflammation and resultant endothelial injury may lead to a hypercoagulable state and predispose patients to micro and macrothrombosis (3, 4).

Lipid modulating agents may limit inflammation and thromboinflammation in COVID-19 by exerting anti-viral, anti-inflammatory, immunomodulatory, and antithrombotic effects (5). Moreover, lower high-density lipoprotein (HDL) cholesterol and higher triglycerides are associated with worse outcomes in patients with COVID-19 (6). Through lipid raft disruption (7), lipid profile improvement, and other effects, lipid modulating agents may impact the outcomes of patients with COVID-19. Moreover, as previously seen in other SARS infections, SARS-CoV-2 infection may lead to the MYD88 gene being highly induced with resultant activation of the nuclear factor kappa-light-chain-enhancer of activated B cells (NF-kB) pathway (8, 9). Statins had inhibitory effects on this pathway, (and a reduction in type 1 interferon) and hyperinflammation (10, 11).

This manuscript summarizes systematically the randomized controlled trials (RCTs) evaluating lipid modulating therapies for the prevention or treatment of COVID-19. The presumed mechanisms of action, existing knowledge of RCTs, as well as knowledge gaps that may influence the design of future trials will be highlighted.

## METHODS

### Data Source and Search Strategy

We searched ClinicalTrials.gov and the World Health Organization International Clinical Trials Registry Platform (WHO-ICTRP) for the identification of RCTs investigating lipid modulating agent trials in COVID-19 (date of last search: March 31, 2021). We used key words for COVID-19 or SARS-CoV-2 or coronavirus disease 2019 and statins (including atorvastatin, rosuvastatin, simvastatin, fluvastatin, lovastatin, pitavastatin, and pravastatin), fibrates (including fenofibrate, clofibrate, bezafibrate, gemfibrozil, and pemafibrate), ezetimibe, bile acid sequestrants (colesevelam, cholestyramine, colestipol), proprotein convertase subtilisin/kexin type 9 (PCSK9) inhibitors (including, alirocumab, evolocumab, and inclisiran), omega-3 fatty acids (including icosapent ethyl, eicosapentaenoic acid [EPA], and docosahexaenoic acid [DHA]), niacin, nicotinic acid, nicotinamide, vitamin B3, evinacumab, mipomersen, lomitapide, bempedoic acid, and cholesteryl ester transfer protein (CETP) inhibitors (anacetrapib, dalcetrapib, evacetrapib, torcetrapib, and TA-8995).

We separated the RCTs in which these agents were used for treatment of patients with COVID-19 versus those used for prevention of the development (or severity) of COVID-19. Study eligibility criteria for inclusion in this review were RCT design with a lipid modifying agent and description of inclusion and exclusion criteria and the primary outcome at clinicaltrials.gov or WHO-ICTRP. Figure 1 describes the search strategy and screening of the studies. For RCTs that met the above eligibility criteria, we searched MEDLINE with PubMed Interface, Google scholar, and pre-print servers including medrxiv.org and biorxiv.org for published design papers or final result manuscripts.

**Figure 1.**
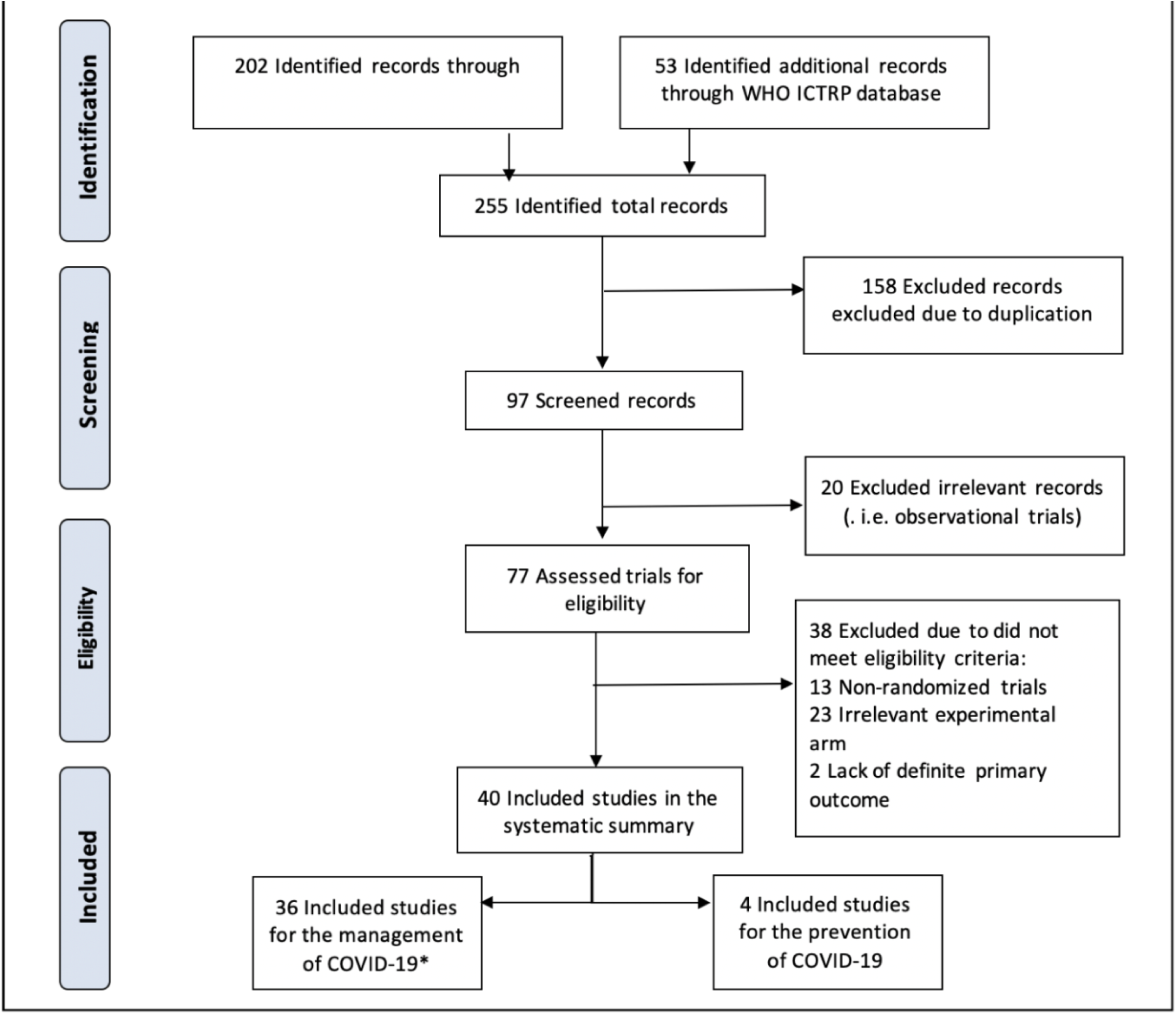
**PRISMA flow diagram.** Caption: Key words used for search of clinicaltrials.gov. and WHO ICTRP included: COVID-19 or SARS-CoV-2 or Coronavirus disease 2019 with statin, atorvastatin, rosuvastatin, simvastatin, fluvastatin, lovastatin, pitavastatin, pravastatin, fibrate, fenofibrate, clofibrate, bezafibrate, gemfibrozil, pemafibrate, ezetimibe, bile acid sequestrant, colesevelam, cholestyramine, colestipol, omega, omega-3, icosapent ethyl, eicosapentaenoic acid, docosahexaenoic acid, PCSK9, PCSK9 inhibitors, Proprotein Convertase Subtilisin/Kexin type 9 alirocumab, evolocumab, inclisiran, niacin, nicotinic acid, nicotinamide, vitamin B3, evinacumab, bempedoic acid, mipomersen, lomitapide, cholesterol ester transfer protein inhibitors, CETP inhibitors, anacetrapib, dalcetrapib, evacetrapib, torcetrapib, and TA-8995. *: One trial identified by discussion with the principal investigator. COVID-19: Coronavirus Disease 2019, PCSK9: Proprotein Convertase Subtilisin/Kexin type 9, WHO ICTRP: World Health Organization International Clinical Trials Registry Platform.

### Summary of the Search Results

We screened 255 records, of which 97 required further manual review. Ultimately, 40 RCTs met the eligibility criteria, of which 36 were related to the management of COVID-19: 17 for statins, 10 for omega-3 fatty acids, 3 for fibrates, 5 for niacin, and 1 for dalcetrapib. In addition, 4 RCTs of omega-3 fatty acids were identified for the prevention of COVID-19. We did not identify RCTs for ezetimibe, bile acid sequestrants, PCSK9 inhibitors, evinacumab, mipomersen, lomitapide, bempedoic acid, or CETP inhibitors other than dalcetrapib for the management or the prevention of COVID-19. A summary of methodological features of the ongoing RCTs categorized by drug class is provided in Table 1. In this table, we describe factors such as number of enrollees, comparator types, blinding, type of primary outcomes (clinical or surrogate outcomes), blinded outcome adjudication, and the existence of published design paper are included. For 7 studies, a design paper or study protocol was available (12–18). Of all RCTs, only 1 has reported the results (at the National Lipid Association Virtual Scientific Sessions) (19).

**Table 1.**
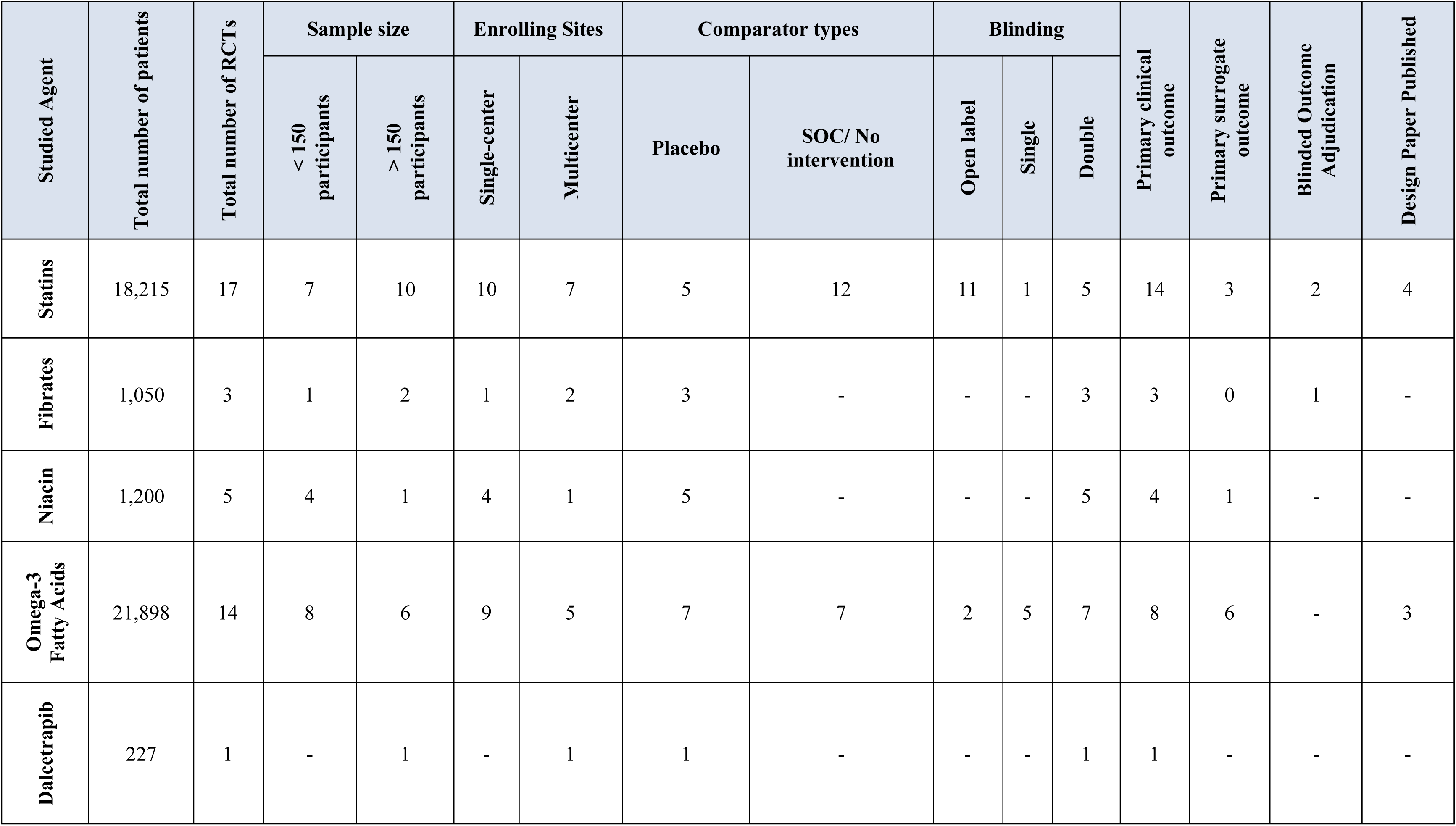
Methodological Features of the Ongoing RCTs Categorized by the Drug Class

### Potential mechanisms of action lipid modulating agents in patients with COVID-19

Figure 2 illustrates the potential pathways through which lipid modulating drugs that have ongoing RCTs may impact the outcomes in COVID-19. These agents include statins, omega-3 fatty acids, fibrates, niacin, and dalcetrapib.

**Figure 2:**
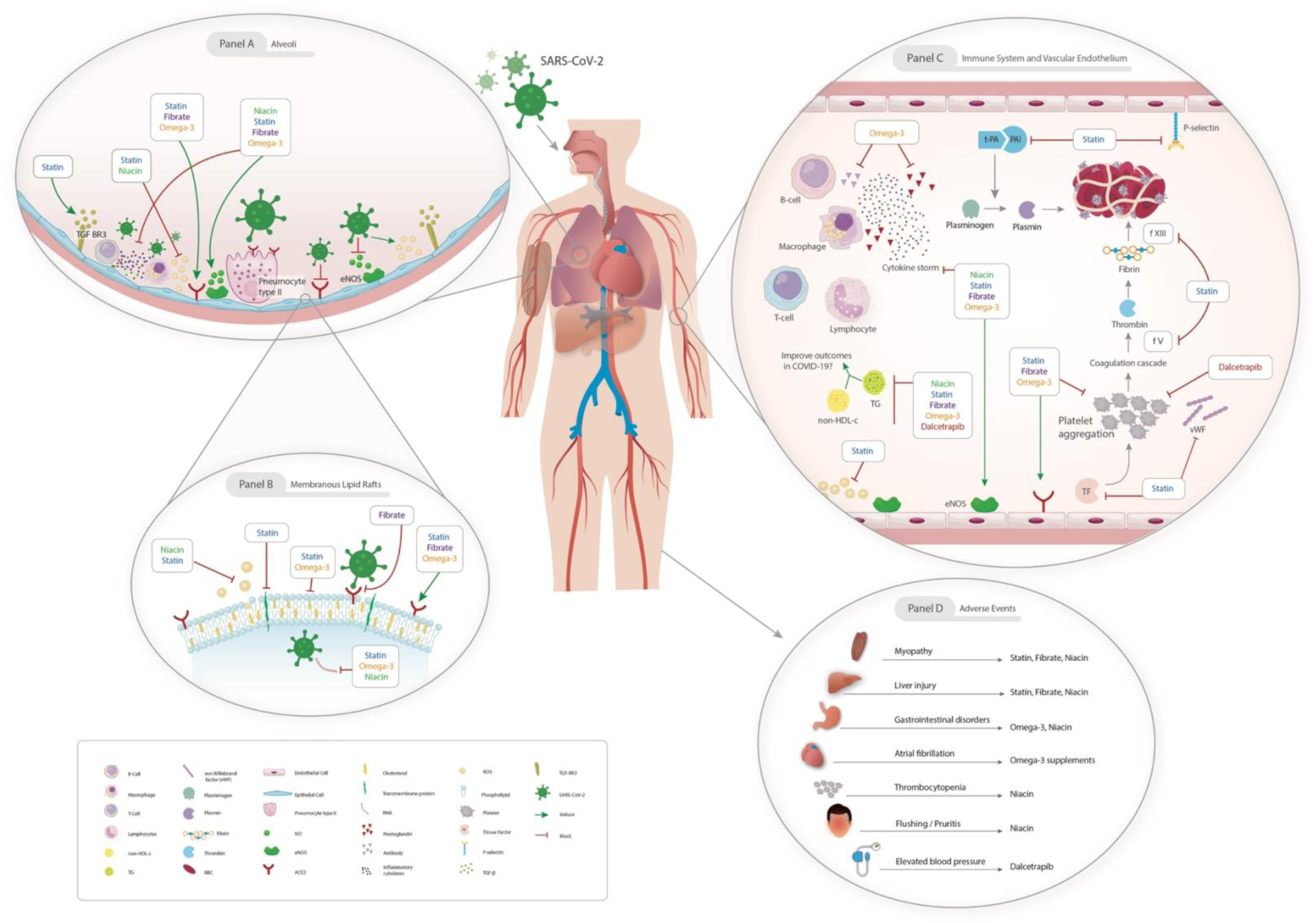
Posited antiviral, immunomodulatory and antithrombotic mechanisms of lipid modulating agents in COVID-19 and their potential adverse effects. **Panel A**: Within the alveoli, the initial binding of the SARS-CoV-2 to the pneumocytes results in innate immune cell (including macrophages and T-cells) infiltration and subsequent cytokine release. Statins, fibrates, omega-3, and niacin may exert immunomodulatory properties to reduce the severity of the inflammatory response. SARS-CoV-2 can downregulate ACE2 expression and reduce its protective effects in many tissues, but statins, fibrates, and omega-3 can maintain the alveoli’s epithelial cell integrity by upregulating ACE2 and increasing the activity of endothelial nitric oxide synthase. Statins may also alleviate collagen deposition and pulmonary fibrosis via upregulation of transforming growth factor beta receptor III. **Panel B**: By destabilized the RBD of the viral spike protein and inhibiting RBD binding to ACE2, fibrates can inhibit viral entry. Statins and omega-3 fatty acids can also inhibit viral entry by disruption in cholesterol synthesis pathway and membranous lipid rafts. There is also some evidence showing the direct anti-viral effects of omega-3 fatty acids on enveloped viruses such as hepatitis C virus by causing leakage and disintegration of virus’ envelope. Statins and niacin can also dampen viral replication via impacts on proteins of the replication pathway. Statin can reduce the prenylation of these proteins and niacin restores nicotinamide-adenine dinucleotide consuming enzyme with anti-viral functions, named as poly-ADP-ribose polymerases. Statin and niacin may have immunomodulatory effects via blocking of the reactive oxygen species. **Panel C**: Within the vessel, lipid modulating agents can modulate the immune response and inhibit the cytokine storm. Omega-3 fatty acids can inhibit the antibody release from the B-cells and also prostaglandins. There is also some evidence showing that statins, fibrates, omega-3 fatty acid, and dalcetrapib may have anti-platelet activities. Moreover, statins may have additional antithrombotic properties by inhibiting tissue factor, von Willebrand factor, factors V, XIII, and plasminogen activator inhibitor. Furthermore, high TG and non-HDL cholesterol is associated with worse outcomes; but it is still unclear that whether lowering triglyceride or non-high-density lipoprotein cholesterol can be used as a therapeutic intervention in COVID-19 or not. **Panel D:** The possible adverse effects of lipid modulating agents are illustrated in Panel D. ACE: Angiotensin Converting Enzyme; COVID-19: Coronavirus Disease 2019; HDL: high-density lipoprotein, RBD: receptor binding domain; SARS-CoV-2: Severe Acute Respiratory Syndrome Coronavirus 2, TG: Triglyceride.

Statins inhibit 3-hydroxy-3-methyl-glutaryl-coenzyme A (HMG-CoA) reductase (20) and the production of isoprenoid intermediates that are critical for viral entry, immune signaling, and the inflammatory cascade (21). These agents also induce transcription factors such as Krüppel-like factor-2, limiting inflammation and prothrombotic functions of activated endothelial cells (22). Statins exert antioxidant and anti-apoptotic effects, potentiate the production of nitric oxide (3, 23), and upregulate transforming growth factor beta receptor III, thereby reducing collagen deposition and pulmonary fibrosis (24). Data in observational studies, RCTs, and meta-analyses in patients with sepsis are controversial. Two recent RCTs showed no improvement in acute respiratory distress syndrome (ARDS) versus placebo (25, 26). However, some studies suggest a benefit associated with statin use. (27) Secondary analysis of the HARP-2 trial, suggested potential improved survival in patients with a high inflammatory status (28). In a meta-analysis of cohort studies and RCTs of patients with ARDS, statin use was not associated with reduced mortality in patients with acute lung injury, but correlated with increased ventilator-free days and reduced sequential organ failure assessment scores (29). In COVID-19, a recent single center retrospective study suggested lower adjusted mortality rates in patients with antecedent statin use compared with non-users (30).

Omega-3 polyunsaturated fatty acids act as a precursor to lipid mediators that reduce inflammation and may prove beneficial in the COVID-19 inflammatory response (14). Icosapent ethyl, an ethyl ester of EPA, has demonstrated anti-inflammatory properties (15). Multiple RCTs have evaluated omega-3 polyunsaturated fatty acids in ARDS. Although individual trial results have been mixed, a meta-analysis showed favorable outcomes with regard to ventilator-free days, length of stay in the intensive care unit (ICU), organ failure, and mortality in patients receiving a diet enriched with EPA and gamma-linolenic acid (31, 32).

In vitro studies suggest that fenofibrate, a fibric acid derivative, destabilizes the receptor binding domain of the SARS-CoV-2 spike protein and inhibits receptor binding domain binding to ACE2. This may reduce viral infectivity by up to 70% (33).

Niacin (nicotinic acid, nicotinamide) increases HDL cholesterol and may reduce the inflammatory mediators. Niacin may also possess anti-viral activity through increasing nicotinamide adenine dinucleotide (NAD) - as nicotinamide restores poly-adenosine diphosphate (ADP)-ribose polymerases functions - which inhibit the viral replication and support innate immunity to SARS-CoV-2 (34).

CETP inhibitors (e.g., dalcetrapib) raise HDL cholesterol, which may have anti-inflammatory properties and inhibit platelet activation (35). However, off-target effects should be considered. In the ILLUMINATE (Investigation of Lipid Level Management to Understand its Impact in Atherosclerotic Events) trial, torcetrapib –another CETP inhibitor –had favorable effects on lipids but showed an increase in death due to sepsis, and increased the systolic blood pressure (36).

## SUMMARY OF THE ONGOING RCTS

A graphical summary of design features of all RCTs for statins, omega-3 fatty acid preparations, fibrates, and niacin for the management of COVID-19 is illustrated in Figures 3, 4, 5, and 6, respectively. In addition, the section on RCTs for the management of patients with diagnosed COVID-19 begins with statin therapy RCTs, followed by RCTs of omega-3 fatty acid preparations, fibrates, niacin, and dalcetrapib. This sequence does not describe treatment preference. Figure 7 illustrates a graphical summary of design features of RCTs for the prevention of COVID-19 (only omega-3 fatty acid had such RCTs). In each section, discussion of these trials is provided according to the clinical setting.

**Figure 3.**
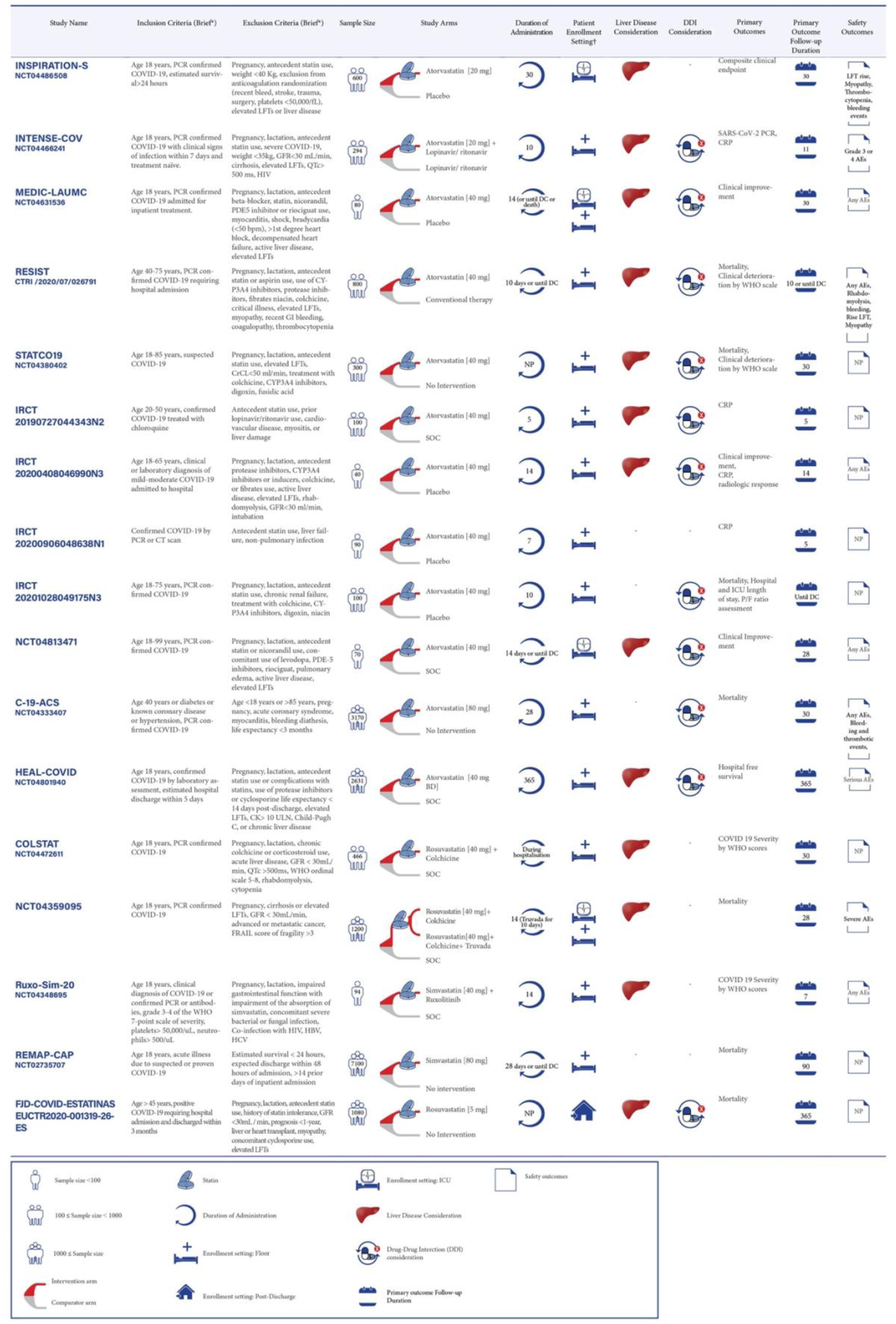
**Ongoing RCTs of statin therapy for Management of Patients with Diagnosed COVID-19**. Caption: Statin therapy trials are evaluating patients in different settings including inpatients (ICU and non-ICU), and post-discharge settings. *: The full list of inclusion and exclusion criteria should be found in the original trial records; ^†^: Patient enrollment setting includes hospitalized ICU or non-ICU patients and post-discharge settings. AEs: Adverse Events; Bpm: Beats per minute; COVID-19: Coronavirus Disease-2019; CK: Creatine Kinase; CrCl: Creatinine Clearance; CRP: C-Reactive Protein; CT: Computed Tomography; DC: Discharge; DDI: Drug-Drug interaction; FRAIL: Fatigue Resistance, Ambulation, Illnesses, and Loss of weight questionnaire; GFR: Glomerular Filtration Rate; GI: Gastrointestinal; QTc: QT Corrected; HBV: Hepatitis B Virus; HCV: Hepatitis C Virus; ICU: Intensive Care Unit; LFT: Liver Function Tests; MI: Myocardial Ischemia; NP: Not Pointed; P/F ratio (PaO2/FiO2) ratio: The ratio of arterial oxygen partial pressure to fractional inspired oxygen; PCR: Polymerase Chain Reaction; PDE5: Phosphodiesterase type 5; RCT: Randomized Controlled Trial; SARS-CoV-2: Severe Acute Respiratory Syndrome-Coronavirus-2; SOC: Standard of Care; ULN: Upper Limit Normal; WHO: World Health Organization.

**Figure 4.**
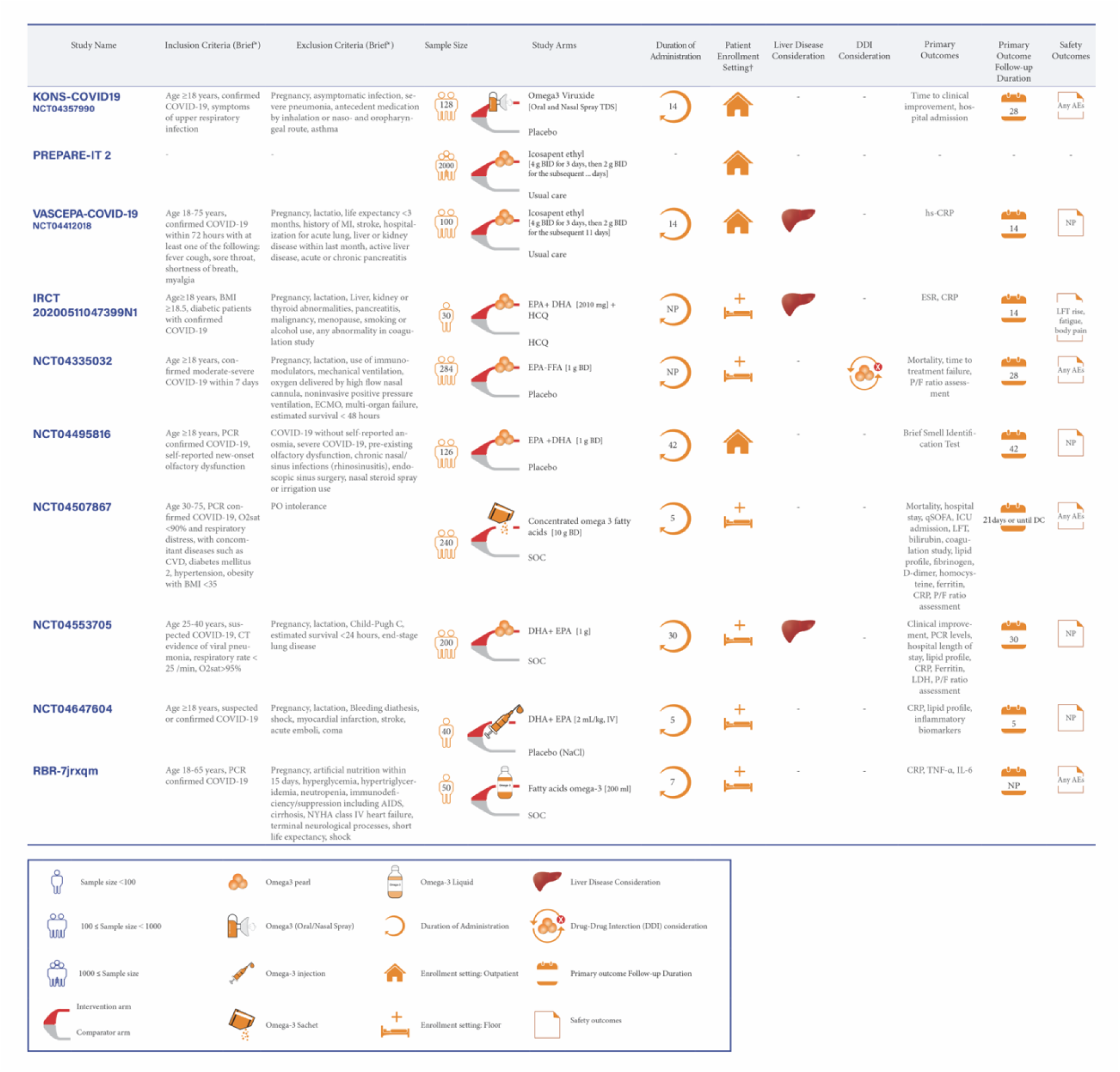
**Ongoing RCTs of Omega-3 Fatty Acid Preparations for Management of Patients with Diagnosed COVID-19**. Caption: Omega-3 fatty acid preparations are evaluating in different settings including inpatients non-ICU, and outpatient settings. *: The full list of inclusion and exclusion criteria should be found in the original trial records; ^†^: Patient enrollment setting includes hospitalized non-ICU patients and outpatient settings. AIDS: Acquired Immunodeficiency Syndrome; BMI: Body Mass Index; CVD: Cardiovascular Disease; DHA: Docosahexaenoic Acid; ESR: Erythrocyte Sedimentation Rate; EPA: Eicosapentaenoic Acid; FFA: Free Fatty Acid; HCQ: Hydroxychloroquine; hs-CRP: high-sensitivity C-Reactive Protein; IL-6: Interleukin-6; IV: Intravenous; LDH: Lactate Dehydrogenase; O_2_sat: Oxygen Saturation; qSOFA: quick Sequential Organ Failure Assessment; TNF-α: Tumor Necrosis Factor-alpha. Other abbreviations as in Figure 3.

**Figure 5.**
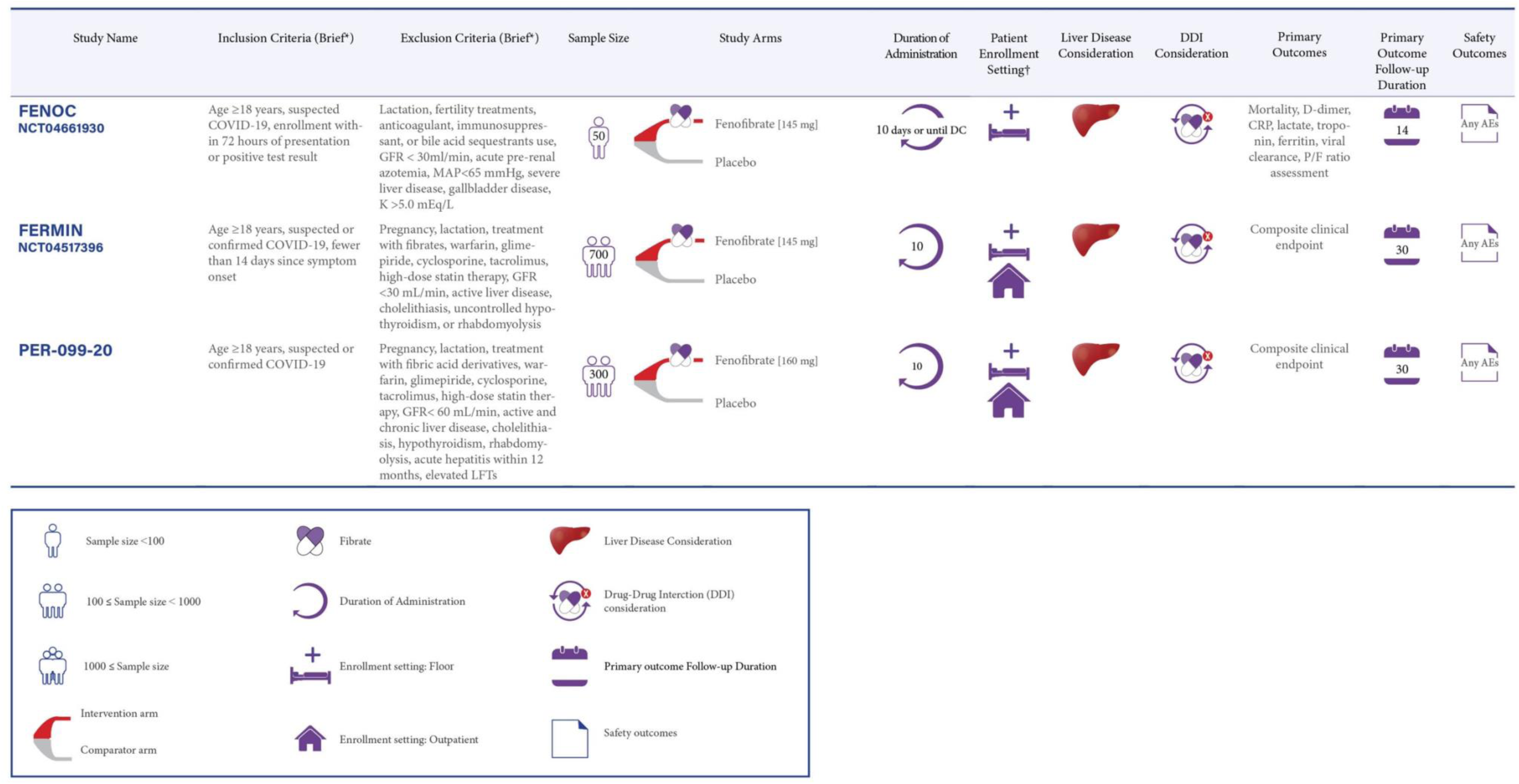
**Ongoing RCTs of fibrates therapy for Management of Patients with Diagnosed COVID-19**. Caption: Fenofibrate is evaluating in different settings including inpatients non-ICU, and outpatient settings. *: The full list of inclusion and exclusion criteria should be found in the original trial records; ^†^: Patient enrollment setting includes hospitalized non-ICU patients and outpatient settings. MAP: Mean Arterial Pressure. Other abbreviations as in Figure 3 and Figure 4.

**Figure 6.**
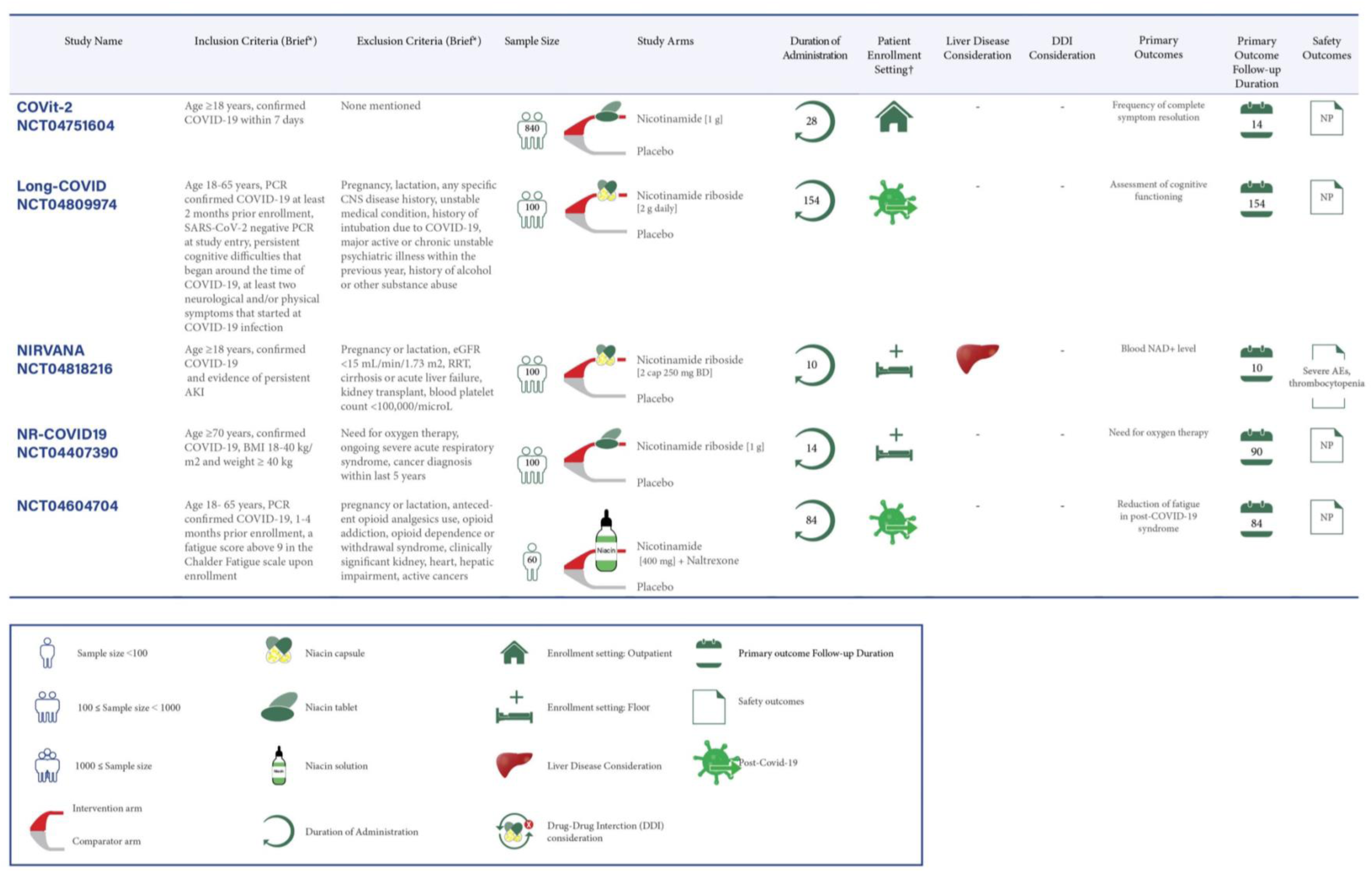
**Ongoing RCTs of niacin therapy for Management of Patients with Diagnosed COVID-19**. Caption: Niacin is evaluating in different settings including inpatients non-ICU, outpatient, and post-acute-COVID-19 settings. *: The full list of inclusion and exclusion criteria should be found in the original trial records; ^†^: Patient enrollment setting includes hospitalized non-ICU patients, outpatient, and post-acute-COVID-19 settings. AKI: Acute Kidney Injury; CNS: Central Nervous System; eGFR: estimated Glomerular Filtration Rate; NAD: Nicotinamide-Adenine Dinucleotide; RRT: Renal Replacement Therapy. Other abbreviations as in Figure 3 and Figure 4

**Figure 7.**
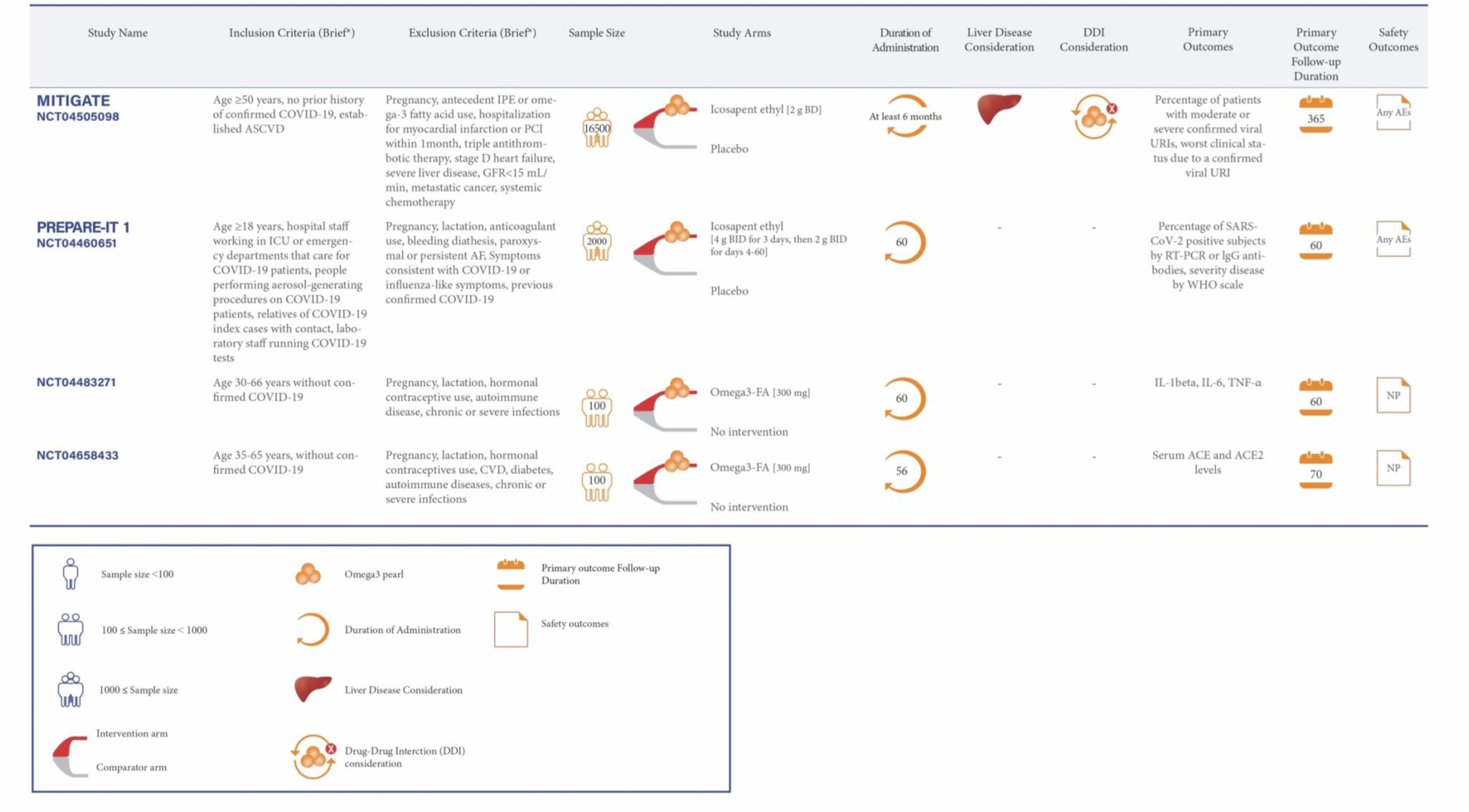
**Ongoing RCTs of Omega-3 Fatty Acid Preparations for Prevention of COVID-19**. Caption: Omega-3 fatty acid preparations are evaluating in moderate to high risk for COVID-19. *: The full list of inclusion and exclusion criteria should be found in the original trial records. ACE: Angiotensin Converting Enzyme; AF: Atrial Fibrillation; ASCVD: Atherosclerotic Cardiovascular Disease; IL-1: Interleukin-1; IPE: Icosapent Ethyl; PCI: Percutaneous Coronary Intervention; RT- PCR: Reverse Transcription Polymerase Chain Reaction; TNF-α: Tumor Necrosis Factor-alpha; URI: Upper Respiratory Infection. Other abbreviations as in Figure 3 and Figure 4.

### RCTs for Management of Patients with Diagnosed COVID-19

#### Ongoing RCTs of statin therapy

Seventeen RCTs of statin therapy have been registered: 16 in the hospitalized setting and 1 in post-discharge setting. These trials assess either moderate-intensity statin therapy (with simvastatin 40 to 80 mg daily or atorvastatin 20 mg daily or rosuvastatin 5 mg daily) or high-intensity statin therapy (with atorvastatin 40 to 80 mg daily or rosuvastatin 40 mg daily) (37).

#### Ongoing RCTs of statin therapy in hospitalized non-ICU patients

Statins are being evaluated in 14 RCTs with the number of participants ranging from 40 to 7,100 patients in the non-ICU hospital settings, including 12 for hospitalized non-ICU patients and 2 that enroll both ICU and non-ICU patients (MEDIC-LAUMC [Managing Endothelial Dysfunction in COVID-19: A Randomized Controlled Trial at LAUMC] and Effectiveness and Safety of Medical Treatment for SARS-CoV-2 (COVID-19) in Colombia [NCT04359095]).

Moderate-intensity statin therapy is being tested in 3 of these 14 RCTs for hospitalized non-ICU patients. The primary outcomes include mortality within 90 days in REMAP-CAP (Randomized, Embedded, Multifactorial Adaptive Platform Trial for Community-Acquired Pneumonia) for 7,100 hospitalized non-ICU patients (17), and assessment of COVID-19 severity by WHO scores within 7 days in the Ruxo-Sim-20 trial (Study of Ruxolitinib Plus Simvastatin in the Prevention and Treatment of Respiratory Failure of COVID-19) for 94 participants. The comparators of these trials are no intervention and standard of care (SOC), respectively. The third RCT of moderate-intensity statin therapy, INTENSE-COV, plans to randomize 294 patients to atorvastatin plus lopinavir/ ritonavir (LPV/r) vs. LPV/r for the co-primary outcomes of the proportions of patients with undetectable SARS-CoV-2 polymerase chain reaction (PCR) and C-reactive protein (CRP) < 27 mg/L at day 11.

High-intensity statin therapy is being assessed in 11 of 14 ongoing RCTs with a total of 8,977 hospitalized non-ICU patients with COVID-19. These 11 RCTs are studying high-intensity statin therapy compared with no treatment (2 of 11) or SOC (5 of 11) or placebo (4 of 11). Mortality during hospitalization or within 10 to 30 days is the most common primary outcome in 5 of 11 RCTs with high-intensity statins for hospitalized non-ICU patients (13, 18). The HEAL-COVID (HElping Alleviate the Longer-term Consequences of COVID-19) trial plans to enroll 2,631 participants to explore the effect of high-intensity statin therapy on hospital free survival within 12 months from enrollment.

Patients with liver disease are excluded in 2 of 3 RCTs with moderate-intensity and 8 of 11 trials with high-intensity statin. Considerations of drug-drug interactions at the time of enrollment were evaluated in 1 of 3 and 8 of 11 RCTs with moderate- and high-intensity statin RCTs.

#### Ongoing RCTs of statin therapy in critically ill patients

Statins are being assessed in 4 RCTs in ICU patients, of which 2 enroll both ICU and non-ICU patients (MEDIC-LAUMC and NCT04359095). Two other RCTs include ICU patients only (INSPIRATION-S [The Intermediate versus Standard-dose Pro-phylactic anticoagulation and Statin In cRitically-ill pATIents with COVID-19: An opeN label randomized controlled trial] with 600 participants (12) and Managing Endothelial Dysfunction in Critically Ill COVID-19 Patients at LAUMCRH [NCT04813471]) with 70 patients with COVID-19.

INSPIRATION-S is the only trial of moderate-intensity statin therapy versus placebo in ICU patients. A composite of all-cause mortality, venous or arterial thrombotic events, and treatment with extracorporeal membrane oxygenation within 30 days is the study primary outcome (12). INSPIRATION-S recently completed patient enrollment and 30-day results are expected in May 2021.

The remaining RCTs in critically ill patients (3 of 4 trials) are investigating high-intensity statin therapy, of which 2 trials (NCT04813471and NCT04359095) are studying high-intensity statin therapy compared with SOC. The MEDIC-LAUMC is assessing the effects of high-intensity atorvastatin versus placebo among 80 participants. In addition, 2 of 3 trials of high-intensity statin therapy (MEDIC-LAUMC and NCT04813471) consider drug-drug interactions before enrollment, and clinical improvement within 1 month is being assessed as the primary outcome in these two trials. The other trial, NCT04359095, assesses the influence of high-intensity statin therapy on mortality within 28 days as the primary outcome among 1,200 ICU or non-ICU patients. Patients with liver disease are excluded in all of the RCTs with moderate or high-intensity statins for critically ill patients.

#### Ongoing RCTs of statin therapy in post-discharge patients

Rosuvastatin (5mg daily) is the only statin-based intervention under investigation in the post-discharge setting. The FJD-COVID-ESTATINAS RCT is evaluating rosuvastatin compared with no treatment in 1,080 patients discharged from hospitalization for COVID19. The primary outcome is a composite of mortality, myocardial infarction, or ischemic stroke within 12 months. Patients with liver disease or concomitant treatment with cyclosporine are excluded. Additional details about ongoing clinical trials of statin therapy are provided in Figure 3.

#### Ongoing RCTs of omega-3 fatty acid preparations

There are 10 ongoing RCTs evaluating the role of omega-3 fatty acid preparations for the management of COVID-19: 6 RCTs in hospitalized non-ICU patients and 4 ongoing RCTs in the outpatient setting.

#### Ongoing RCTs of omega-3 fatty acid preparations in hospitalized non-ICU patients

Omega-3 fatty acids are being evaluated in 6 RCTs with the number of participants ranging from 30 to 284 patients in the non-ICU hospital settings. Most RCTs (5 of 6) assess the oral use of omega-3 fatty acid preparations compared with SOC (3 of 5) or with placebo (1 of 5). The oral use of omega-3 fatty acid preparations plus hydroxychloroquine compared with hydroxychloroquine alone is assessed in the Comparison of the effectiveness of omega-3 and Hydroxychloroquine on Inflammatory factors, liver enzymes, and clinical symptoms in diabetic COVID-19 patients (IRCT20200511047399N1). Only one RCT (NCT04647604 [Resolving Inflammatory Storm in COVID-19 Patients by Omega-3 Polyunsaturated Fatty Acids]) explores intravenous administration of omega-3 fatty acid preparations versus placebo (14). Moreover, 3 of 6 RCTs of hospitalized non-ICU patients have co-primary outcomes including inflammatory markers and lipid levels. Elevated liver enzymes are being evaluated as the safety outcome only in the IRCT20200511047399N1 trial. Patients with liver disease are excluded in 2 of the 6 RCTs of omega-3 fatty acid in hospitalized non-ICU patients.

#### Ongoing RCTs of omega-3 fatty acid preparations in outpatient setting

Omega-3 fatty acid preparations are being evaluated in 4 ongoing RCTs for treatment of COVID-19: KONS-COVID19 (Viruxal Oral and Nasal Spray for Treating the Symptoms of COVID-19), VASCEPA-COVID-19 (An Investigation on the Effects of Icosapent Ethyl (Vascepa^TM^) on Inflammatory Biomarkers in Individuals With COVID-1), PREPARE-IT 2 (Prevention of COVID19 With EPA in Healthcare Providers at Risk - Intervention Trial 2) and COVID-19 Anosmia Study (NCT04495816) (16).

The KONS-COVID19 trial tests omega-3 inhaled use versus placebo among 128 outpatient participants. The primary outcome is time to clinical improvement within 28 days. The VASCEPA-COVID-19 trial assesses the oral use of icosapent ethyl compared with usual care in a total of 100 outpatient participants. High sensitivity CRP (hsCRP) level is the primary outcome in VASCEPA-COVID-19. Patients with active severe liver disease are excluded in this trial. Initial findings from the VASCEPA-COVID-19 were presented at National Lipid Association Scientific Sessions, and showed that the use of icosapent ethyl for 14 days reduced hs-CRP: 3.2 vs. 1.6 mg/L (p = 0.011) and led to symptom improvement assessed by InFLUenza Patient-Reported Outcome (FLU-PRO) score in outpatients with COVID-19 after 14 days (19). The PREPARE-IT 2 trial is assessing the effects of icosapent ethyl versus placebo in 2,000 outpatient participants. The impact of oral use of omega-3 fatty acid preparations versus placebo on olfactory performance within 6 weeks among outpatients is under evaluation in the NCT04495816 trial with 126 participants. Additional details about ongoing RCTs of omega-3 fatty acid preparations for treatment of COVID-19 are described in Figure 4.

#### Ongoing RCTs of fibrates

There are three RCTs investigating fibrates in patients with COVID-19. These RCTs are testing fenofibrate versus placebo in hospitalized non-ICU patients: FENOC (Fenofibrate for Patients With COVID-19 Requiring Hospitalization), FERMIN (FEnofibRate as a Metabolic INtervention for COVID-19) and Fenofibrate as A Metabolic Intervention for Coronavirus Disease 2019 (COVID-19): A Randomized Controlled Trial (PER-099-20). FERMIN and PER-099-20 also enrolled outpatients. The sample size of these studies ranges from 50 to 700 patients.

The main outcomes in the FENOC include improvement in laboratory markers, the ratio of arterial oxygen partial pressure to fractional inspired oxygen (PaO_2_/FiO_2_), and mortality. The composite endpoint as the primary outcome for FERMIN and PER-099-20 trials will be a global rank score that grades patients based on survival, need for respiratory/mechanical support, the fraction of inspired oxygen/ percent oxygen saturation (FiO_2_/ SpO_2_), the number of days out of the hospital for outpatient participants who are hospitalized after enrollment, and the modified borg dyspnea scale for the outpatient subset not hospitalized. All these trials consider drug-drug interactions prior to the enrollment. Patients with active liver disease are excluded in all 3 of these trials. Additional information about these RCTs is summarized in Figure 5.

#### Ongoing RCTs of niacin

Five RCTs of niacin therapy were identified: 2 in hospitalized non-ICU patients, 1 in outpatient setting, and 2 ongoing RCTs in post-acute COVID-19. All of these trials are studying niacin compared with placebo.

#### Ongoing RCTs of niacin in hospitalized non-ICU patients

Niacin is being assessed in 2 ongoing RCTs for 100 hospitalized non-ICU patients in each trial: NIRVANA (Nicotinamide Riboside in SARS-CoV-2 (COVID-19) Patients for Renal Protection) and NR-COVID19 (Effects of Nicotinamide Riboside on the Clinical Outcome of Covid-19 in the Elderly) trials. The primary outcomes of these trials are the alteration of blood NAD+ level within 10 days and need for oxygen therapy with a follow-up duration of 90 days, respectively. In NIRVANA, thrombocytopenia is being evaluated as the safety outcome, and patients with liver disease are being excluded.

#### Ongoing RCTs of niacin in outpatient setting

The COVit-2 (Improvement of the Nutritional Status Regarding Nicotinamide (Vitamin B3) and the Disease Course of COVID-19) trial is the only trial of niacin therapy versus placebo in outpatient setting with 840 patients planned to be enrolled. The frequency of complete symptom resolution within 2 weeks is the primary outcome.

#### Ongoing RCTs of niacin in post-COVID-19 setting

Niacin is being evaluated in 2 RCTs in the post-COVID-19 setting: Long-COVID (Clinical Trial of Niagen to Examine Recovery in People with Persistent Cognitive and Physical Symptoms After COVID-19 Illness) and Pilot Study Into LDN and NAD+ for Treatment of Patients With Post-COVID-19 Syndrome (NCT04604704) trials. These RCTs assess the oral use of niacin in 100 participants and iontophoresis patches for 60 patients with COVID-19, respectively.

The Long-COVID trial is studying the impact of niacin on the cognitive function of patients with positive PCR at least 2 months prior to enrollment. The primary outcome follow-up duration is 22 weeks. NCT04604704 plans to enroll 60 patients with PCR-confirmed COVID-19 1 to 4 months prior to enrollment. This trial assesses the reduction of fatigue in post-COVID-19 syndrome within 12 weeks as the primary outcome. Patients with liver disease are excluded in the NCT04604704 trial. Additional details about RCTs of niacin are illustrated in Figure 6.

#### Ongoing RCT of CETP inhibitors

The dal-COVID (Effect of Dalcetrapib in Patients with Confirmed Mild to Moderate COVID-19) trial is the only trial of dalcetrapib (900mg, 1,800mg, and 3,600mg daily) versus placebo in 208 outpatients with mild-to-moderate COVID-19. The primary outcome is time to sustained symptom resolution within 28 days. Patients with liver disease are excluded from NCT04676867 trial. Drug-drug interactions are being considered before enrollment.

### RCTs for Prevention of Contracting COVID-19

Use of omega-3 fatty acid preparations as a preventive measure against COVID-19 is being investigated in 4 RCTs: MITIGATE (A Pragmatic Randomized Trial of Icosapent Ethyl for High-Cardiovascular Risk Adults) (15), PREPARE-IT 1 (Prevention of COVID19 With EPA in Healthcare Providers at Risk - Intervention Trial 1), The Effect of Omega-3 on Selected Cytokines Involved in Cytokine Storm (NCT04483271), and The Effect of Omega-3 Supplements on the Serum Levels of ACE/ACE2 Ratio as a Potential Key in Cardiovascular Disease and COVID-19 (NCT04658433).

MITIGATE and PREPARE-IT 1 trials are studying the effects of icosapent ethyl versus placebo. In turn, NCT04483271 and NCT04658433 assess omega-3 fatty acid supplements compared with no treatment for 100 patients in each trial. The number of confirmed viral infections and worst clinical status due to a viral upper respiratory infection are the co-primary outcomes for MITIGATE with 16,500 participants. The number of confirmed viral infections is the primary outcome in PREPARE-IT 1 with 2,000 participants. The primary outcomes for the NCT04483271 and NCT04658433 trials are inflammatory markers such as interleukin-1beta (IL-1β), IL-6, tumor necrosis factor-alpha, and serum ACE and ACE2. Patients with severe liver disease are excluded only in MITIGATE. Additional details about these RCTs are summarized in Figure 7.

## DISCUSSION

The perspective of the COVID-19 disease state has broadened from pneumonia to a systemic multi-organ disease, with systemic inflammation and thrombosis as key features (2, 38). The present review identified 34 RCTs that evaluate the role of lipid modulating agents in the management of acute COVID-19, 2 RCTs in patients with post-acute COVID-19, and 4 RCTs for prevention of contracting (or severity of) COVID-19. Results from these trials may expand the armamentarium for management of COVID-19. The neutral results of recent RCTs of escalated-dose anticoagulation in critically-ill patients with COVID-1 (39, 40) may indicate the significance of the maladaptive immune response in severe COVID-19 (41). It is in this context that lipid modulating agents with pleotropic effects offer possible therapeutic potential (42). The moderate immunomodulating effect of these agents lessens the chance of excessive immunosuppression and superinfection, commonly seen with other anti-inflammatory agents.

Despite such hope, certain methodological limitations of some of the ongoing RCTs deserve attention. These include small sample size, use of primary surrogate outcomes, and lack of blinded outcome adjudication, which hamper the rigor of the trials. More than one year into the pandemic, 40 RCTs of lipid modifying therapies were identified, with only 21 (with total estimated sample size of 7,675) having double blind design. The results from only one trial have been communicated in preliminary form. In addition, despite the scientific rationale summarized in this manuscript, translation to clinical benefit is not assured. Among the immune-modulating therapies, only steroids have shown consistent efficacy in patients with COVID-19 (43–47). The neutral results with ivermectin (48) and hydroxychloroquine (49), and the mixed results with colchicine (50, 51) and tocilizumab (52, 53), remind us that biological plausibility may not translate to meaningful treatment. Hence, the ongoing lipid modulating therapy RCTs are of particular interest.

### Statins as Multi-Purpose Drugs?

Despite initial concern that statins might increase expression of ACE2 and facilitate SARS-CoV-2 entry with potential deleterious effects, observational studies suggest an association between antecedent statin use and improved survival. Lee et al. in a population-based propensity matched study of 10,448 patients with COVID-19, showed a significant reduction in hazard of death in statin users compared with non-users (hazard ratio: 0.64 [95% CI, 0.43–0.95]; P=0.02) (54). Gupta et al. in a retrospective propensity matching analysis of 1,296 patients with COVID-19 reported similar reduction in the odds of death with antecedent statin use (OR: 0.47, 95% CI 0.36–0.62, p < 0.001) (30). Such non-randomized observational studies are subject to confounding, including confounding by indication, and need confirmation in ongoing RCTs. There are several limitations to the ongoing statin investigations. Several RCTs do not include thrombotic events among their pre-specified outcomes. Quality of life is evaluated in only 3 RCTs. The role of statin therapy in post-discharge setting for patients with COVID-19 is being studied in only 1 RCT with 1080 patients, but not in the outpatient setting.

### Omega-3 Polyunsaturated Fatty Acids: Hope or Hype?

Anti-inflammatory effects (55) and potential impact on ARDS progression (56) make omega-3 fatty acids worthwhile agents for investigation. Functional limitations and quality of life are evaluated exclusively in outpatient trials, while mortality and clinical improvement are evaluated in hospitalized patients. Two trials with considerable sample size evaluate the role of icosapent ethyl as in prevention of contracting COVID-19.

Several limitations apply to the omega-3 fatty acid RCTs: The total number of patients enrolled in omega-3 fatty acids trials for management of COVID-19 is relatively few (Figure 4). Heterogeneity in use of various formulations (EPA, DHA, or EPA-DHA; ethyl esters vs. free fatty acids)and impurities, and/or oxidative alterations in unregulated supplement preparations makes it more difficult to determine the specific effects of each. Icosapent ethyl will be studied in two large outpatient studies (PREPARE-IT 1 and MITIGATE) trials as a preventive agent, but not in the inpatient setting.

### Fibrates: New Spark for a Dying Candle?

Despite the declining use for cardiovascular risk reduction (57), fibrates may decrease viral entry and SARS-CoV-2 infectivity by increasing sulfatide levels (58) and inhibiting the receptor binding domain to ACE2 (33). Strengths of ongoing fibrate trials include endpoint selection: death, ARDS-related outcomes, inflammatory markers and invasive mechanical support are the primary outcome under evaluation. Drug interactions were generally considered with fenofibrate initiation. However, fenofibrate is the only fibrate under investigation in RCTs including patients with COVID-19, and its lack of benefit in cardiovascular disease prevention may raise skepticism regarding its utility in COVID-19.

### Niacin: Time for a Comeback?

Anti-inflammatory effects (34) and potential protection against lung injury (59) have made niacin a target for investigation in COVID-19. Niacin is under evaluation in both acute and post-acute settings. Clinical improvement and symptom resolution are primary outcomes in the majority of RCTs. RCTs in the post-COVID-19 setting will evaluate fatigue and cognitive function for 3 to 6 months.

As with fenofibrate, lack of benefit and presence of adverse effects with nicotinamide seen in prior cardiovascular RCTs (60, 61) may dampen excitement for use in COVID-19. Trials in the inpatient and post-acute setting are limited by small sample size and variability in niacin dosing may lead to heterogenous results.

### CETP Inhibitors: Room for Potential Benefit?

Low HDL is associated with increased severity of COVID-19 and correlates with approved biomarkers such as ferritin and D-dimer levels (6). Dalcetrapib will be the first CETP inhibitor to be tested in patients with mild to moderate COVID-19 with time to symptom resolution as the primary outcome. Of note, dalcetrapib did not improve clinical outcomes in those with acute coronary syndromes (62). Among CETP inhibitors, only anacetrapib showed a modest cardiovascular benefit (9% relative risk reduction) (63), whereas torcetrapib (64, 65) led to increase in systolic blood pressure and worse cardiovascular outcomes. Findings from dal-COVID will help clarify whether dalcetrapib merits further testing in COVID-19.

### Expecting the Unexpected: Possible Adverse Effects for Lipid Modulating Agent Trials

Based on the knowledge from RCTs in cardiovascular diseases, important adverse effects should be monitored when the results of these RCTs accrue. Myopathy is the most common adverse effect of statin use and is managed by conversion to other statin formulations. Severe muscle complications (i.e., rhabdomyolysis) are exceedingly rare (66). Rise in liver enzymes is another potential but infrequent adverse effect. Fibrates may also increase the risk of myopathy and hepatocyte injury (67). Drug-drug interactions play an important role in this increased risk, and is considered in the ongoing COVID-19 RCTs (68). Other risks associated with statin use including hemorrhagic stroke in those with previous stroke are not of clinically important magnitude (69–71). New-onset or worsening of atrial fibrillation is a concern with omega-3 fatty acids (72–74) and increased bleeding due to the effects on platelet aggregation should be monitored in the RCT results.

The adverse effects of niacin include flushing, pruritus, gastrointestinal disorder, thrombocytopenia, hyperuricemia, hyperglycemia, myopathy, and hepatotoxicity (75). Only the NIRVANA trial assesses thrombocytopenia as a safety outcome and excluded patients with liver disease.

These potential risks are limited by the short duration of treatment in most trials. Of note, the adverse events in massive event-driven cardiovascular trials result from multiple months to years of treatment, whereas the current COVID-19 trials generally have much shorter treatment duration. The possible adverse effects of lipid modulating agents are illustrated in Figure 2.

## CONCLUSIONS

Lipid modulating agents may mitigate the multiorgan damage associated with COVID-19 through anti-inflammatory, anti-viral, and pleiotropic effects. Findings from ongoing rigorously conducted and adequately powered RCTs can assess the possible efficacy of lipid modulating agents in the prevention or treatment of various stages of COVID-19, and may open new horizons for research and clinical practice.

## Data Availability

Data availability only after permission of corresponding author

## ABBREVIATIONS AND ACRONYMS

ACE2: angiotensin converting enzyme 2
ARDS: acute respiratory distress syndrome
CETP: cholesterol ester transfer protein
COVID-19: coronavirus disease 2019
HDL: high-density lipoprotein
HMG-CoA: 3-hydroxy-3-methyl-glutaryl-coenzyme A
ICU: intensive care unit
NAD: nicotinamide adenine dinucleotide
RCT: randomized controlled trial
SARS-CoV-2: severe acute respiratory syndrome coronavirus 2

## ACKNOWLEDGEMENTS

The authors would like to express their sincere gratitude to Fatemeh Esmaeili, MS, for her kind assistance in the preparation of the Figures.

## HIGHLIGHTS

- Lipid modulating agents have pleiotropic effects, including anti-viral, immunomodulatory, and antithrombotic properties that might help treat COVID-19.
- 36 RCTs are evaluating the impact of lipid modulating agents including statins, omega-3, fibrates, niacin, and CETP inhibitors for treatment of COVID-19.
- Omega-3 fatty acid preparations are being investigated for prevention of COVID-19 in 4 ongoing RCTs.
- Niacin is being assessed in 2 RCTs for post-acute-COVID-19.

---

**Central Illustration: Summary of ongoing lipid modulating agents’ trials in COVID-19.** Caption: Statins are the most frequently studied lipid modulating agents in RCTs of patients with COVID-19. Omega-3 fatty acid preparations are the only studied lipid modulating agents in prevention of COVID-19. Niacin is the only lipid modulating agent being studied for post-acute COVID-19. For more details please review Figure 3, 4 and 5. COVID-19: Coronavirus Disease 2019, FA: Fatty Acid, ICU: Intensive Care Unit, IPE: Icosapent ethyl, PCSK9: Proprotein Convertase Subtilisin/Kexin type 9.

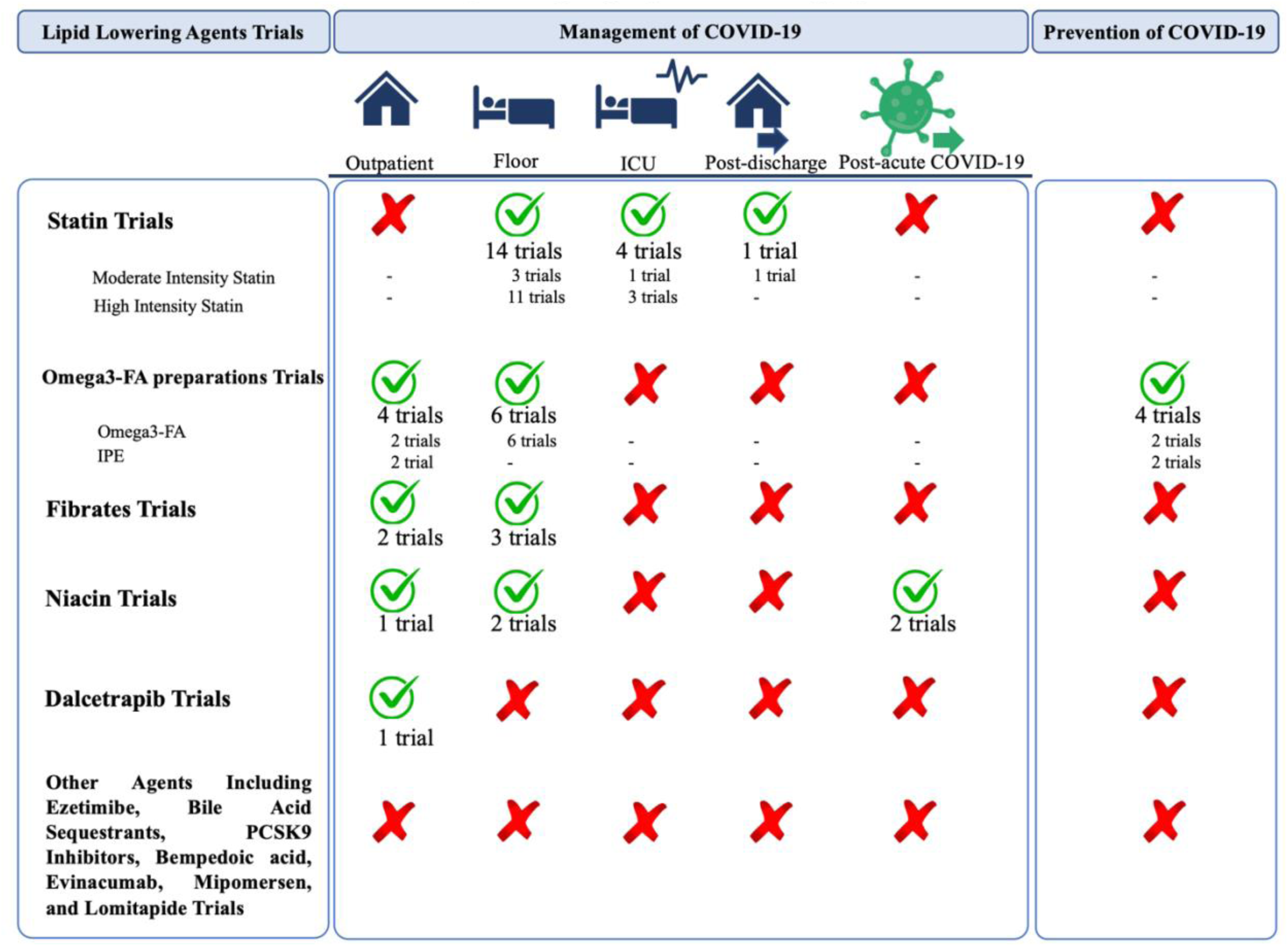

